# Attitudes Toward The COVID-19 Vaccine Among North Carolina Participants In The COVID-19 Community Research Partnership

**DOI:** 10.1101/2021.05.20.21257343

**Authors:** Chukwunyelu H Enwezor, James E. Peacock, Sharon L Edelstein, Amy N Hinkelman, Austin L Seals, Thomas F Wierzba, Iqra Munawar, Patrick D Maguire, William H Lagarde, Michael S Runyon, Michael A Gibbs, Thomas R Gallaher, John W. Sanders, David M. Herrington

## Abstract

Willingness to receive the newly developed Coronavirus Disease-2019 (COVID-19) vaccines is highly variable. To assess the receptiveness of a select sample of North Carolinians to COVID-19 vaccination, a brief survey was conducted among participants in the COVID-19 Community Research Partnership (CCRP) affiliated with five medical centers in North Carolina. A total of 20,232 CCRP participants completed a multiple choice, mini survey electronically between December 17, 2020 and January 13, 2021. Of the 20,232 survey respondents, 15,422 (76.2%) were receptive to vaccination. Vaccine receptiveness increased incrementally with age with those >70 years being the most willing to be vaccinated compared to all other age groups. Respondents with no previous COVID-19 diagnosis were more likely to accept the vaccine compared to those that have a previous COVID-19 diagnosis (76.6% vs 60.9%). Comparative analysis of gender, race/ethnicity, and residence locale revealed that women, African Americans, and suburban participants were less willing to get a COVID-19 vaccine. There was no difference in vaccine intent based on healthcare worker status. Of those unwilling to get the vaccine, 82% indicated that the reason was uncertainty about the safety and efficacy of the vaccine.

## Introduction

Since cases of COVID-19 were first reported in Wuhan, China in December 2019, various measures like mask wearing and social distancing have been instituted by various countries (1,2,3). Despite these measures, there have been significant loss of lives and livelihoods. A lot of effort was put into developing vaccines as the most effective means of stopping the virus. By April 2021, three COVID-19 vaccines had been approved for Emergency Use Authorization (EUA) by the Food and Drug Administration (FDA). About 200 million vaccines had been administered nationwide (6) and 6.7 million in the state of North Carolina (**Error! Reference source not found.**) as of April 22, 2021. Despite the initial enthusiasm about vaccine availability, vaccine acceptance and willingness to undergo vaccination were not universal with some national surveys suggesting that only 50-55% of respondents would be willing to receive the COVID-19 vaccine (**Error! Reference source not found.**,6). In North Carolina, some polls revealed intent at only 40-45% (8).

To assess the extent of vaccine hesitancy in North Carolina, we conducted a survey between December 17, 2020 and January 13, 2021 of 20,232 individuals affiliated with five medical centers from differing geographic regions of North Carolina. This report presents the key findings from that survey.

## Materials and Methods

The CCRP is a multi-site, prospective study combining daily electronic symptom surveillance, longitudinal serologic surveillance, and electronic health record capture. Demographic and survey data was collected via a secure, HIPAA-compliant, online portal. The study has received approval by centralized IRB (Wake Forest Baptist Health). Five of the sites participated in this sub-study. Those sites were Campbell University in Buies Creek, NC, New Hanover Regional Medical Center in Wilmington, NC, Wake Forest Baptist Health in Winston-Salem, NC, WakeMed Health and Hospitals in Raleigh, NC, and Vidant Health, in Greenville, NC.

Basic demographic data captured for all CCRP participants included age, sex, previous COVID-19 diagnosis status, community of residence, and race/ethnicity. Counties of residence were characterized as urban, suburban, or rural utilizing the North Carolina rural center counties map. Densities were calculated based on the 2014 census population estimates (9). Participants were also classified as to whether they were healthcare workers with all healthcare-related vocations and disciplines included in that category. In addition to their daily CCRP surveys, participants were asked to complete a single multiple-choice mini-survey on attitudes about COVID-19 vaccination ^1^. There were four choices for vaccine intent: yes, no, undecided, and prefer not to answer. For participants who did not respond “yes” to the vaccine intent question, a follow-up question asked participants to specify reasons for vaccine hesitancy (Fig 1). The survey was sent daily from December 17, 2020 until January 13, 2021. Once a response was documented electronically, the mini survey was deleted from further daily surveys to ensure that each participant responded only once.

**Fig 1.**
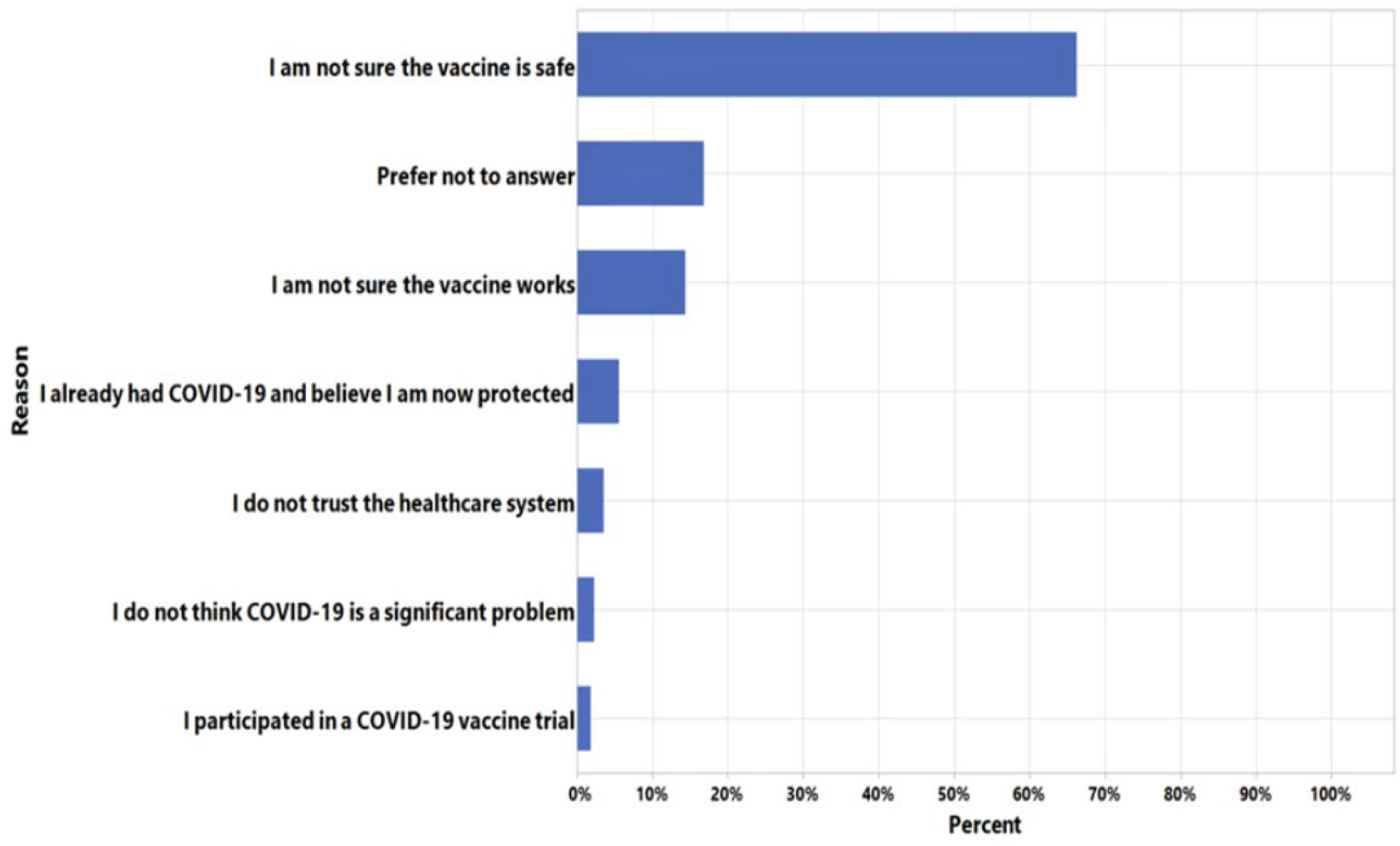
Reasons given by participants that did not respond “yes” on the survey.

To correlate the association of demographic characteristics with vaccine intent, binomial regression was implemented. Vaccine intent responses were categorized into two groups for this analysis: Yes and No/Undecided. To assess how demographic characteristics were associated with vaccine receptiveness, multivariate binomial regression was used with a log link function. The resulting coefficient estimates were exponentiated (e^b^) to calculate relative risk. The relative risk of responding ‘Yes’ was calculated for each demographic variable, along with 95% confidence intervals. Previous COVID-19 Diagnosis was self-reported information included as a part of the enrollment questionnaire. The impact of a prior diagnosis of COVID-19 on vaccine receptiveness was also analyzed in a similar fashion. P-values<0.001 were considered significant. All statistical analysis was conducted using SAS 9.4 (SAS Institute, Cary, NC).

## Results

A total of 20,232 people completed the mini survey (Campbell University, n^2^=147; New Hanover Regional Medical Center, n=641; Wake Forest Baptist Health, n=16058; Wake-Med Health and Hospitals, n=2419; and Vidant Health, n=967). Most respondents were White (90%), female (68%), and most were <60 years old (median age 50 years with range 18 to 80). Non-healthcare workers comprised 74% of the total respondents. Only 2.4% (n=476) of participants had a previous COVID-19 diagnosis. Across all demographics, 76.2% of participants expressed intent to get the vaccine (Table 1).

**Table 1.** Vaccine intent responses grouped by sex, age, race & ethnicity, healthcare worker status, previous COVID-19 diagnosis, Site and county class. n- no; P – prefer not to answer; N – no; Y - yes.

|  | Total | Vaccine Intent |  |  |  |
| --- | --- | --- | --- | --- | --- |
|  |  | No/Undecided/P |  | Yes |  |
|  | n | n | Percent | n | Percent |
| **TOTAL** | 20232 | 4810 | 23.8 | 15422 | 76.2 |
| **SEX** |  |  |  |  |  |
| Women | 13784 | 3688 | 26.8 | 10096 | 73.2 |
| Men | 6448 | 1122 | 17.4 | 5326 | 82.6 |
| **AGE GROUP** |  |  |  |  |  |
| <30 | 1096 | 322 | 29.4 | 774 | 70.6 |
| 30-39 | 3086 | 889 | 28.8 | 2197 | 71.2 |
| 40-49 | 3897 | 1175 | 30.2 | 2722 | 69.8 |
| 50-59 | 4256 | 1180 | 27.7 | 3076 | 72.3 |
| 60-69 | 4898 | 865 | 17.7 | 4033 | 82.3 |
| 70+ | 2999 | 379 | 12.6 | 2620 | 87.4 |
| **ETHNICITY** |  |  |  |  |  |
| Black or African American | 932 | 426 | 45.7 | 506 | 54.3 |
| Hispanic or Latino | 413 | 122 | 29.5 | 291 | 70.5 |
| Other | 758 | 179 | 23.6 | 579 | 76.4 |
| White (not Hispanic/Latino) | 18129 | 4083 | 22.5 | 14046 | 77.5 |
| **HEALTHCARE WORKER** |  |  |  |  |  |
| N | 15062 | 3466 | 23 | 11596 | 77 |
| Y | 5170 | 1344 | 26 | 3826 | 74 |
| **PREVIOUS COVID-19 DIAGNOSIS** |  |  |  |  |  |
| N | 19756 | 4624 | 23.4 | 15132 | 76.6 |
| Y | 476 | 186 | 39.1 | 290 | 60.9 |
| **SITE** |  |  |  |  |  |
| Campbell University | 147 | 48 | 32.7 | 99 | 67.3 |
| New Hanover Regional Medical Center | 641 | 100 | 15.6 | 541 | 84.4 |
| Vidant Health | 967 | 285 | 29.5 | 682 | 70.5 |
| Wake Forest Baptist Health | 16058 | 3917 | 24.4 | 12141 | 75.6 |
| WakeMed Health and Hospitals | 2419 | 460 | 19 | 1959 | 81 |
| **COUNTY CLASS** |  |  |  |  |  |
| Rural | 6645 | 1916 | 28.8 | 4729 | 71.2 |
| Suburban | 1669 | 530 | 31.8 | 1139 | 68.2 |
|  | 11918 | 2364 | 19.8 | 9554 | 80.2 |

Compared to men, women were less likely to indicate a willingness to undergo vaccination [82.6% vs 73.2% adjusted RR=0.92, 95% CI (0.91,0.93), P<0.0001] (Table 2). Vaccine receptiveness increased incrementally with age, with those >70 years the most likely to accept the vaccine compared to each of the other age categories (Table 1).

**Table 2.**
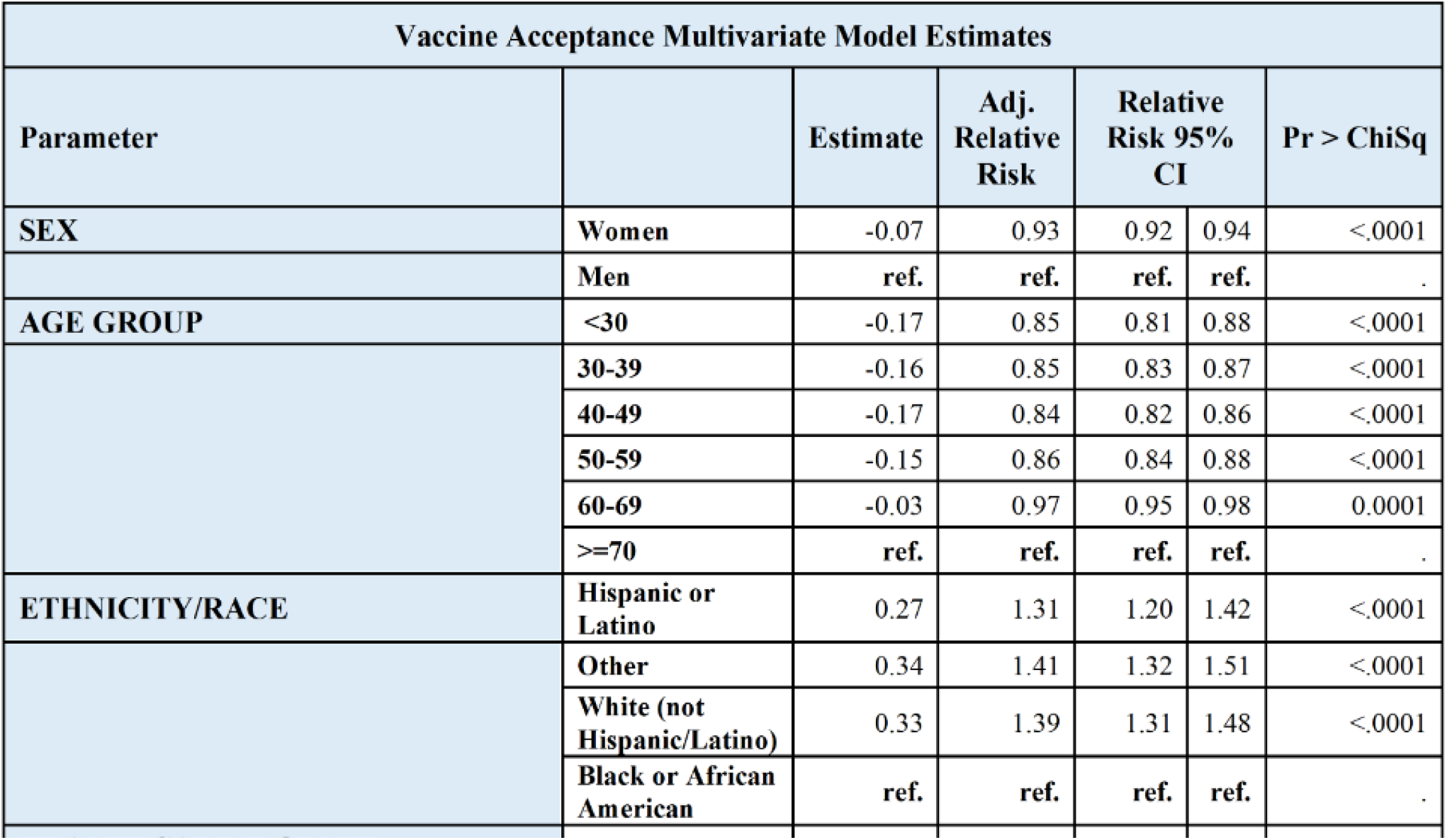
Vaccine acceptance multivariate and type 3 analysis grouped by sex, age, race & ethnicity, healthcare worker status, previous COVID-19 diagnosis, Site and county class. n- no; P– prefer not to answer; N – no; Y – yes; CI – Confidence interval; ChiSq – Chi square

| Vaccine Acceptance Multivariate Model Estimates |  |  |  |  |  |  |
| --- | --- | --- | --- | --- | --- | --- |
| Parameter |  | Estimate | Adj. Relative Risk | Relative Risk 95% CI |  | Pr > ChiSq |
| SEX | Women | -0.07 | 0.93 | 0.92 | 0.94 | <.0001 |
|  | Men | ref. | ref. | ref. | ref. | . |
| AGE GROUP | <30 | -0.17 | 0.85 | 0.81 | 0.88 | <.0001 |
|  | 30-39 | -0.16 | 0.85 | 0.83 | 0.87 | <.0001 |
|  | 40-49 | -0.17 | 0.84 | 0.82 | 0.86 | <.0001 |
|  | 50-59 | -0.15 | 0.86 | 0.84 | 0.88 | <.0001 |
|  | 60-69 | -0.03 | 0.97 | 0.95 | 0.98 | 0.0001 |
|  | >=70 | ref. | ref. | ref. | ref. | . |
| ETHNICITY/RACE | Hispanic or Latino | 0.27 | 1.31 | 1.20 | 1.42 | <.0001 |
|  | Other | 0.34 | 1.41 | 1.32 | 1.51 | <.0001 |
|  | White (not Hispanic/Latino) | 0.33 | 1.39 | 1.31 | 1.48 | <.0001 |
|  | Black or African American | ref. | ref. | ref. | ref. | . |

Vaccine Acceptance Multivariate Model Estimates
| Parameter |  | Estimate | Adj. Relative Risk | Relative Risk 95% CI |  | Pr > ChiSq |
| --- | --- | --- | --- | --- | --- | --- |
| SEX | Women | -0.07 | 0.93 | 0.92 | 0.94 | <.0001 |
|  | Men | ref. | ref. | ref. | ref. | . |
| AGE GROUP | <30 | -0.17 | 0.85 | 0.81 | 0.88 | <.0001 |
|  | 30-39 | -0.16 | 0.85 | 0.83 | 0.87 | <.0001 |
|  | 40-49 | -0.17 | 0.84 | 0.82 | 0.86 | <.0001 |
|  | 50-59 | -0.15 | 0.86 | 0.84 | 0.88 | <.0001 |
|  | 60-69 | -0.03 | 0.97 | 0.95 | 0.98 | 0.0001 |
|  | >=70 | ref. | ref. | ref. | ref. | . |
| ETHNICITY/RACE | Hispanic or Latino | 0.27 | 1.31 | 1.20 | 1.42 | <.0001 |
|  | Other | 0.34 | 1.41 | 1.32 | 1.51 | <.0001 |
|  | White (not Hispanic/Latino) | 0.33 | 1.39 | 1.31 | 1.48 | <.0001 |
|  | Black or African American | ref. | ref. | ref. | ref. | . |
| HEALTHCARE WORKER | N | -0.03 | 0.97 | 0.96 | 0.99 | 0.0014 |
|  | Y | ref. | ref. | ref. | ref. | . |
| PREVIOUS COVID-19 DIAGNOSIS | N | 0.18 | 1.20 | 1.11 | 1.28 | <.0001 |
|  | Y | ref. | ref. | ref. | ref. | . |
| COUNTY CLASS | Rural | -0.10 | 0.90 | 0.89 | 0.92 | <.0001 |
|  | Suburban | -0.16 | 0.85 | 0.83 | 0.88 | <.0001 |
|  | Urban | ref. | ref. | ref. | ref. | . |

African Americans were the least willing to accept the vaccine at 54.3%, compared to all other groups; Hispanic or Latino (70.5%), other (76.4%) and White (77.5%) (Adjusted RR 1.31,1.41 and 1.39 respectively p<0.0001)

Respondents without a previous COVID-19 diagnosis were more likely to accept the vaccine compared to those that have a previous COVID-19 diagnosis (76.6% vs 60.9%, adjusted RR 1.20, 95% CI (1.11,1.28), p<0.0001)

When stratifying response by county residence, suburban and rural residents were less likely than urban residents to accept the vaccine. Suburban residents were the least likely to accept the vaccine (68%) as compared to urban residents (81%) (Adjusted RR 0.85, 95% CI (0.83,0.88), p<0.0001). The vaccine acceptance rate of rural residents was 71.2%.

Interestingly, healthcare workers were somewhat less likely to accept the vaccine compared to non-healthcare workers [74% vs 77%] (Table 1), a difference that remained statistically significant in a multivariate analysis (p=0.0014) (Table 2).

Among the 4,810 individuals who indicated that they did not intend to get vaccinated or were undecided, the most common reasons were concerns about safety (67%) and efficacy (15%) (Fig 1). An important point to note is that of the participants that did not respond “yes” to the vaccine, more than half were undecided^3^.

## Discussion

This survey was designed to assess attitudes about vaccine acceptance among a cohort of North Carolinians during the period immediately following EUA of the Pfizer and Moderna COVID-19 vaccines. Most participants in this study expressed willingness to receive a vaccine.

Previously published surveys have addressed vaccine hesitancy (6,7,12,**Error! Reference source not found**.,13,13). The largest of these was a global study done in April 2020 involving 13,426 participants from 19 countries, including 773 participants in the US. Of the US participants, 75.2% of the respondents reported intent to get the vaccine (10,13). A survey conducted by the CDC between September and December of 2020 showed 68% of the participants reporting their intent to undergo vaccination (11). A survey done by the Kaiser Family Foundation (KFF) in February 2021, found only 55% of respondents intending to get the vaccine (6). Another recent poll conducted February 16-21 by Pew Research was headlined *“growing share of Americans say plan to get a COVID-19 vaccine or already have”*. In this survey, 69% of the public intend to get the vaccine, or had already received the vaccine. This finding compared to a similar poll conducted by Pew in November 2020, is up by 19% (12). Although the questions asked in these surveys were not strictly comparable to ours, multiple surveys indicate between 50%-75% of respondents report an intent or likely intent to get the vaccine.

Looking at the subgroups in our survey, the strongest independent predictor of COVID-19 vaccine hesitancy is Black race. This is particularly important to note because racial and ethnic minority groups, have disproportionately been affected by COVID-19. Despite Black Americans making up just 12.5% of the US population, an October 2020 CDC report showed that 18.7% of COVID-19 cases were Black Americans (14). In another report, it was noted that the mortality rate among Black Americans is 2 -fold higher than Whites (15). In Black American communities and other minority populations, mistrust of the healthcare system due to inequities in access and prior adverse experiences with medical research has been the main reason for hesitancy (16,17). On local, state and national levels, there have been various interventions aimed at achieving an equitable vaccine campaign and decreasing hesitancy in Black, Indigenous and People of Color (BIPOC) communities in the United States. Grassroot community partnerships and educational programs like “train-the-trainer” led by the COVID-19 Prevention Network (CoVPN) of the National Institute of Allergy and Infectious Diseases (NIAID) and on a national level, guidance provided by the current administration based on 5 key principles: iteration, involvement, information, investment and integration, may have helped decrease hesitancy in the Black American community since our survey was conducted(18,19). Recent surveys show a trend in the Black community towards vaccine acceptance (20,21). On the other hand, resident locale has now become a stronger predictor of vaccine hesitancy with residents of rural counties least likely to accept the COVID-19 vaccine (21,23,23,25). This shift may be related to political and religious affiliations (25,27). In our survey, it was interesting to note that suburban residents were the least likely to accept the vaccine (68.2%), compared to rural (71.2%) and urban residents (80.2%). While many reports of vaccine hesitancy among rural residents nationally have been noted, this is the first survey that shows more hesitancy among suburban residents. North Carolina rural residents are significantly more accepting of the COVID-19 vaccine when compared to national data from a KFF survey in January 2021, which showed only 31% of rural respondents had intent to get the COVID-19 vaccine (23).

Other independent predictors of hesitancy to get the COVID-19 vaccine in our survey are younger adults, female gender and having a previous COVID-19 diagnosis. Among younger adults, misconceptions about risk for adverse outcomes from COVID-19 infection and uncertainty of vaccine effectiveness may be the primary barriers to vaccine acceptance (26). Women are less likely to accept the vaccine in the survey. Other surveys show that safety and efficacy of the vaccines are the primary reasons for hesitancy (27). The implications of this could potentially be significant because studies have shown that women make approximately 80% of the healthcare decisions for their families (28). Participants with previous COVID-19 diagnosis are less likely to accept the vaccine because they believe they are protected (Fig 1). A study looking at immune memory generated after COVID-19 infection showed retained immunity at ∼6 months (29). This immunity wanes over time. Hence, the CDC has recommended COVID-19 vaccination be offered to individuals that have recovered from the infection following good safety data from clinical trials Finally, for all groups, poor availability of scheduling information, lack of internet access, inability to take time off from work, and/or transportation difficulties which preclude travel to vaccination sites may also need to be addressed (31).

An interesting group in this survey are healthcare workers. Healthcare workers in this survey include, all healthcare related vocations and disciplines, clinical staff members, support staff and pharmacies. Healthcare workers trended towards not accepting the vaccine when compared to non-healthcare worker (74% v 77%). Although, this goes contrary to what we should expect, similar attitudes have been reflected in other surveys (32,33). This is relevant because healthcare workers can transmit confidence in the vaccine to the communities that they serve thereby increasing vaccine uptake.

Among participants in the survey that did not indicate an intent to get the COVID-19 vaccine, more than half (56%) of the respondents were undecided because of uncertainty of efficacy and safety. This informs us that there is an opportunity to address the concerns this group of individuals have. The CDC “Vaccinate with Confidence” national strategy (34) provides a framework for public health officials and advocacy groups to reinforce confidence in vaccines:

- Build trust.
- Empower healthcare personnel.
- Engage communities and individuals.

These strategies should be targeted to the underlying reasons for vaccine hesitancy specific to groups undecided or unwilling to pursue vaccination. A “before we attempt to persuade, try to understand” approach may yield better success in communities less willing to be vaccinated (34).

There are several potential limitations of this study. First, our data represent responses from respondents in North Carolina enrolled into an ongoing research study through regional healthcare systems and may not be representative of national data. Our volunteers are likely to be better connected to a healthcare system and more comfortable with electronic communication than the general community. Second, this survey provides a snapshot in time during the early rollout of the vaccines, and attitudes may have changed with more recent nationwide and local interventions to address hesitancy. Third, the demographics of our survey participants may not reflect national demographics. Fourth, our finding of more hesitancy among suburban residents may simply reflect limitations in our method for characterizing counties as it is difficult to accurately describe many counties in North Carolina as entirely urban, suburban, or rural. Finally, survey results might not be comparable to other national or state polls or surveys due to potential differences in survey methods, sample population, and questions related to willingness to get the vaccine.

In conclusion, in this survey of participants in North Carolina from mid-December 2020 to January 2021, a large majority indicated intent to undergo COVID-19 vaccination. Our survey results suggest that vaccination hesitancy may be a potential impediment to a successful vaccination campaign, especially among several subgroups. Opportunities exist to target those subgroups with focused educational interventions and other support in the hope of increasing vaccine acceptance. Further work is planned to assess changing attitudes about COVID-19 vaccination.

## Supporting information

Questionnaire and Supplemental table

## Data Availability

NA

## Abbreviations

BIPOC: Blacks, Indigenous and People of color
CCRP: COVID-19 Community Research Partnership
CDC: Centers for Disease Control and Prevention
COVID-19: Coronavirus disease 2019
CoVPN: COVID-19 Prevention Network
EUA: Emergency Use Authorization
FDA: Food and Drug Administration
KFF: Kaiser Family Foundation
NC: North Carolina
NIAID: National Institute of Allergy and Infectious Diseases
SARS-CoV-2: severe acute respiratory syndrome coronavirus-2
US: United States
WHO: World Health Organization

## Author Contributions

C.E. served as lead author. A.S contributed to the analysis of the results J.P., J.S., D.H., A.H, S.E., T.W., I.M., P.M., M.G., M.R., W.L., S.H., all contributed to the design and implementation of the research, and to the writing of the manuscript.

All authors have read and approved the final manuscript.

## Funding

This work was supported by the Coronavirus Aid, Relief, and Economic Security (CARES) Act by the U.S. Department of the Treasury.

## Institutional Review Board Statement

The study was conducted according to the guidelines of the Declaration of Helsinki and approved by the Institutional Review Board of Wake Forest School of Medicine Institutional Review Board, protocol [IRB00064912].

## Informed Consent Statement

Informed consent was obtained from all subjects involved in the study.

## Data Availability Statement

The datasets used and/or analyzed during the current study are available from the corresponding author upon request.

## Acknowledgements

**COVID-19 Community Research Partnership Study Group:**

**Wake Forest School of Medicine**

Mark A. Espeland PhD, Morgana Mongraw-Chaffin PhD, Alain Bertoni MD, Martha A. Alexander-Miller PhD, Allison Mathews PhD, MS, Brian Ostasiewski, Christine Ann Pittman Ballard MPH

**George Washington Biostatistics Center**

Diane Uschner PhD, Michele Santacatterina PhD, Greg Strylewicz PhD, Brian Burke MS, Mihili Gunaratne MPH, Meghan Turney MA, Shirley Qin Zhou MS

**Atrium Health**

, MPH, Lewis H. McCurdy MD, Yhenneko Taylor PhD, Lydia Calamari MD, Hazel Tapp PhD, Amina Ahmed MD, Michael Brennan DDS, Lindsay Munn PhD, RN, Tim Hetherington MS, Lauren Lu, Connell Dunn, Melanie Hogg MS, CCRA, Andrea Price, Mariana Leonidas, Laura Staton, Kennisha Spencer MPH, Melinda Manning, Whitney Rossman MS, Frank Gohs MS, Anna Harris MPH, Bella Gutnik MS, Jennifer Priem PhD, MA

**MedStar**

Kristen Miller DrPH, CPPS, William Weintraub MD, Chris Washington, Allison Moses, Sarahfaye Dolman, Julissa Zelaya-Portillo, John Erkus, Joseph Blumenthal, Romero Barrientos, Ronald E, Sonita Bennett, Shrenik Shah, Shrey Mathur, Christian Boxley, Paul Kolm, Long La, Cheng Zhang, Eva Hochberger, Ella Franklin, Deliya Wesley, Naheed Ahmed

**University of Maryland School of Medicine - Baltimore**

Karen Kotloff MD, Wilbur Chen MD, MS, DeAnna Friedman-Klabanoff MD, Andrea Berry MD, Helen Powell, PhD

**Tulane University**

Joseph Keating PhD, Patricia Kissinger PhD, Richard Oberhelman MD, John Schieffelin MD, Joshua Yukich PhD, Andrew “AJ” Beron MPH, Devin Hayes BS, Johanna Teigen MPH

**U of Mississippi**

Adolfo Correa MD, PhD, Leandro Mena MD, MPH, Bhagyashri Navalkele MD, Yuan-I Min MD, Alexandra Castillo MPH, Lori Ward PhD, MS, Robert P. Santos MD, Courtney Gomillia MS-PHS, Pramod Anugu, Yan Gao MPH, Jason Green, Ramona Sandlin RHIA, Donald Moore MS, Lemichal Drake, Dorothy Horton RN

**WakeMed Health and Hospitals**, LaMonica Daniel BSCR

New Hanover Regional Medical Center, Lynette McFayden, RN

**Vidant Health**

Thomas Gallaher, MD, Michael Zimmer, PhD Danielle Oliver, Tina Dixon

**Campbell University**

, Robin King-Thiele DO, Terri S. Hamrick PhD, Chika Okafor MD, Regina B. Bray Brown MD, Pinoorma Vinod MD

## Conflicts of Interest

The authors declare no conflict of interest. The funders had no role in the design of the study; in the collection, analyses, or interpretation of data; in the writing of the manuscript, or in the decision to publish the results.

## Footnotes

1 See supplemental data for questionnaire.

2 n - Number of participants

3 Supplemental data. Table 3

## Notes

### Competing Interest Statement

The authors have declared no competing interest.

### Author Declarations

Institutional Review Board Statement: The study was conducted according to the guidelines of the Declaration of Helsinki and approved by the Institutional Review Board of Wake Forest School of Medicine Institutional Review Board, protocol [IRB00064912].

