## Supplementary material for "Attitudes Toward The COVID-19 Vaccine Among North Carolina Participants In The COVID-19 Community Research Partnership": Questionnaire and Supplemental table

SUPPLEMENTAL INFORMATION

QUESTIONNAIRE

**COVID-19 Vaccine Survey**

Q1. Have you already been vaccinated against COVID-19, or do you intend to be vaccinated once you are eligible to receive it?

- Yes
- No
- Unsure
- Prefer not to answer.

Q2. If you responded “No” or “Unsure” please indicate why (select all that apply)

a. I already had COVID-19 and believe I am now protected.
b. I received the active vaccine in a COVID-19 vaccine trial.
c. I do not think COVID-19 is a significant problem.
d. I am not sure the vaccine works.
e. I am not sure the vaccine is safe.
f. I don’t trust the healthcare system.
g. Prefer not to answer.

Questionnaire sent to participants of the survey.

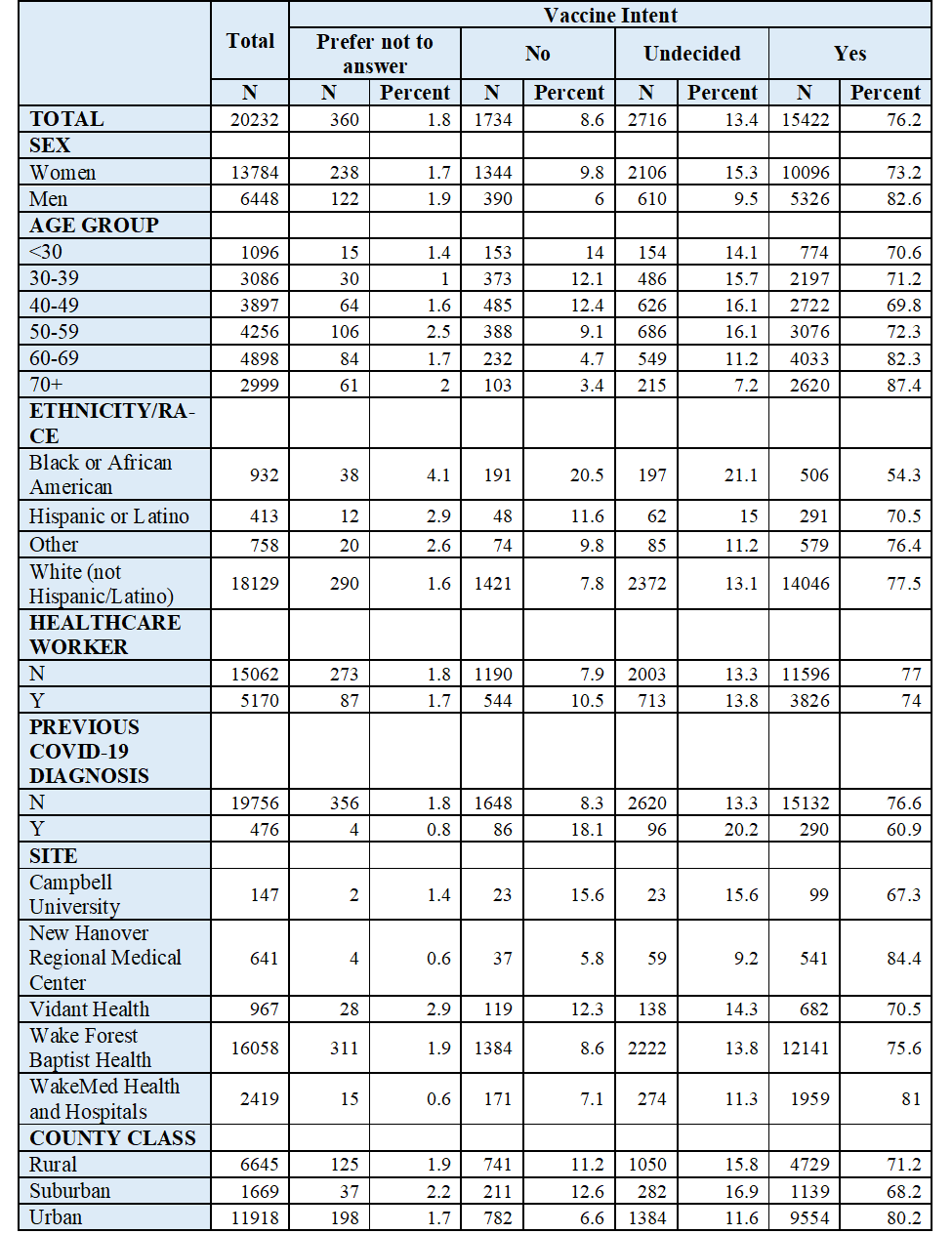

Table 3. Breaking the “no” group to prefer not to answer, no and undecided.
